# Examining the Factor Structure of the DSM-5 Level 1 Cross-Cutting Symptom Measure

**DOI:** 10.1101/2021.04.28.21256253

**Authors:** Alison Gibbons, Cristan Farmer, Jacob S Shaw, Joyce Y Chung

**Author notes:** **Corresponding author:** Joyce Y Chung, MD, National Institute of Mental Health, NIH Clinical Center, 10 Center Dr., Bldg. 10, Rm 6-5340, Bethesda, MD 20892. **Disclosures:** The authors report no financial relationships with commercial interests.

## Abstract

**Objectives:** The DSM-5 Level 1 Cross-Cutting Symptom Measure (DSM-XC) is a transdiagnostic mental health symptom measure that has shown promise in informing clinical diagnostic evaluations and as a screening tool for research. However, few studies have assessed the latent dimensionality of the DSM-XC. We examined the factor structure of the DSM-XC in a large convenience sample of participants with varying degrees of psychological health.

**Methods:** Participants (n=3533) enrolled in a protocol conducted at the National Institute of Mental Health (NCT04339790). We used a factor analytic framework to evaluate an existing two-factor solution (Lace & Merz, 2020) and two additional candidate solutions.

**Results:** The Lace and Merz solution had acceptable fit. Exploratory factor analysis yielded two candidate solutions: a six-factor (characterized as mood, worry, activation, somatic, thoughts, and substance use) and a bifactor (general factor of non-specific psychopathology, residual factors characterized as internalizing and thought disorder), which both had good fit and full measurement invariance across age, sex, and enrollment date.

**Conclusions:** Our findings confirm that the DSM-XC may be conceptualized as a multidimensional instrument and provide a scoring solution for researchers who wish to measure distinct constructs. Future research on the psychometric profile of the DSM-XC is needed, focused on the validity of these candidate solutions and their performance across research populations and settings.

## 1. Introduction

According to the 2019 National Survey on Drug Use and Health (NSDUH) by the Substance Abuse and Mental Health Services Administration (SAMHSA), an estimated 20.6%, or 51.5 million adults aged 18 or older in the United States were diagnosed with any mental illness (AMI), defined as a mental, behavioral, or emotional disorder that can vary in impairment ((1)). Furthermore, fewer than half (44.8%) of those with AMI received mental health services in the past year. Given these high rates of current mental illness and low rates of treatment, there is a need to better assess mental health problems in the general population. While there are existing mental health measures in use, many are limited to symptoms within one diagnostic category, such as the Patient Health Questionnaire 9 (PHQ-9), which focuses on depression ((2)). In addition, because symptoms associated with discrete mental health diagnoses often co-occur, a tool that is broader and transdiagnostic would be useful both in clinical settings and in research studies.

The DSM-5 Level 1 Cross-Cutting Symptom Measures (DSM-XC) were developed by the American Psychiatric Association (APA) as transdiagnostic mental health symptom measures (adult, child and parent versions). They were developed in response to recommendations that the DSM-5 include dimensional assessments in addition to categorical diagnoses utilized in previous versions of the DSM (3). The adult measure includes 23 self-report questions covering 13 cross-diagnostic domains of psychopathology: depression, anger, mania, anxiety, somatic symptoms, suicidal ideation, psychosis, sleep problems, memory, repetitive thoughts and behaviors, dissociations, personality functioning, and substance use (4). The survey was intended to inform a clinical diagnostic evaluation and provide clinicians with a tool to track the presence, frequency, and severity of cross-cutting psychiatric symptoms (5). An early study evaluated the measure in routine clinical practice and found it to be both clinically useful and favorably viewed by clinicians and patients (6). It has also been recommended for use in clinical research (7).

In clinical practice, the DSM-XC is to be used as a Level 1 screener for mental illness followed by Level 2 measures of a specific domain depending on initial responses. The limited psychometric data available for the DSM-XC primarily pertain to this purpose. Our group used a modified version of the DSM-XC to screen volunteers for mental health research and found good specificity but poor sensitivity when used alone, but improved sensitivity when combined with additional clinical data, e.g., use of psychotropic medication (8). Conversely, among patients with substance use disorders in a correctional program, the DSM-XC had good sensitivity but poor specificity for DSM diagnoses of depression, mania, psychosis, and anxiety (9).

Though most reports on the DSM-XC described its use in clinical practice, it has also been recommended for use as a dimensional measure of psychopathology in research studies. Of note, the DSM-XC is one of the measures in the minimal list of data collection measures required by the National Institute of Mental Health (NIMH) for its grantees who conduct mental health human subjects’ research. The minimal list has been identified by NIMH, the Wellcome Trust and other funders of mental health research using the PhenX consensus process (https://www.phenxtoolkit.org/collections/view/1) or the International Consortium for Health Outcomes Measurement (ICHOC) (https://www.ichom.org/resource-library/category/conditionspecific-resources/depression-anxiety). Given that the need for such a measure was the impetus behind the DSM-XC’s development, it is surprising that few published studies have assessed its latent dimensionality or provided guidance on how to interpret DSM-XC responses at the item, domain or full survey level. One study that administered the DSM-XC to non-treatment seeking college students found it to have acceptable convergent validity when the domain total scores were compared to longer measures of the intended mental health construct (10). A study conducted by the APA in concert with DSM field trials administered the survey to adult psychiatric patients and reported mean scores for the DSM-XC items in their supplementary materials. However, they made no claims that the scores within domains measured a unidimensional construct. Whether and how the items included in the DSM-XC measure dimensional mental health constructs (e.g., depression) is an empirical question that should be further investigated.

Given the transdiagnostic design of the DSM-XC survey, the latent dimensionality of the measure should be explored. To date, three studies have published on the factor structure of the DSM-XC, each with limitations. A Turkish study reported an exploratory factor analysis (EFA) on data collected from healthy volunteers and psychiatric patients (n=206) and found that a three-factor structure capturing neurosis, psychosis, and substance use best suited the data (11). Using confirmatory factor analysis (CFA), another group demonstrated adequate fit of the 13 DSM-XC domain structure to data from 362 Pakistani prisoners (12). Both studies used translated versions of the measure (Turkish and Urdu, respectively) and collected data from distinct populations (i.e., psychiatric patients and their relatives, prisoners), which could limit the generalizability of their findings. A recent study employed both EFA and CFA methods on the adult version of the DSM-XC in English (13). They recruited and collected data from 400 participants using Amazon Mechanical Turk, a crowdsourcing website (n=191 EFA, n= 209 CFA). Lace and Merz report that a two-factor model, comprising externalizing/serious mental illness and internalizing/affective constructs, best fit their data. All three studies utilized sample sizes which were small relative to recommendations by psychometric (14) and none performed invariance analyses to demonstrate robustness of the identified solution. Notably, none of the three studies described utilized a bifactor approach to analyzing their data. This was the approach described in the seminal paper by Caspi and colleagues in which they propose a general psychopathology factor (p factor) which was the most parsimonious explanation of psychiatric disorders in a large longitudinal data set (15).

In this report, we sought to address the limitations outlined above by examining the factor structure of the DSM-XC in a large sample of participants enrolled in a protocol conducted through the National Institute of Mental Health Intramural Research Program (NIMH IRP) (ClinicalTrials.gov identifier: NCT04339790). This online study was launched to examine the mental health impact of the COVID-19 pandemic among a convenience sample with a wide range of mental health histories, and the psychometric evaluation of the DSM-XC is a secondary analysis. Following an EFA to identify candidate solutions, we employed CFA to evaluate the fit of the two-factor solution proposed by Lace and Merz (13), the fit of our proposed solutions, and the measurement invariance of our proposed solutions across age, sex, and enrollment date.

## 2. Method

These data were collected as part of a NIMH study that was carried out in accordance with the latest version of the Declaration of Helsinki and was approved by the Institutional Review Board of the National Institutes of Health. For this purpose of this report, we will primarily focus on methodology relevant to the collection of the DSM-XC survey data.

### 2.1 Participants

Participants were recruited through a variety of means, including email invitations, social media ads, list-serv postings, clinicaltrials.gov and direct mail postcards. Exclusion criteria were minimal: participants were required to be 18 years or older and to speak and understand English. The sample was intended to be representative of the general population. For this analysis, we included participants who had provided responses to all 22 DSM-XC items at the time of enrollment (n=3533).

### 2.2 Measure

The DSM-XC was chosen for the parent study because of its cross-diagnostic approach to mental health symptoms. In addition, the DSM-XC was promoted by the NIMH as a common measure in funded research projects (NOT-MH-15-009, 2015). The measure consists of 23 items, each rated on a 5-point Likert scale (0=none – not at all, 4=severe – nearly every day). We chose to omit one item on thoughts of self-harm (question 11) because the study was conducted online and therefore participants who may endorse self-harm could not be reliably contacted for further safety evaluation.

### 2.3 Procedures

Study enrollment occurred from April 4^th^ to November 15^th^, 2020. After signing an electronic consent form, participants used a secure website to complete several surveys, including the DSM-XC. Participants were then invited to complete a smaller set of surveys which included the DSM-XC every two weeks for 6 months. The study website was HIPAA compliant and did not collect personally identifiable information. Compensation was not provided for participation.

### 2.4 Statistical Analysis

#### 2.4.1 Data preparation

The analysis set of cross-sectional DSM-XC responses of 3,533 individuals at time of enrollment was separated into exploratory (n = 1,202) and confirmatory sets (n = 2,331) using a randomly generated number for each participant. Response category cell size for each item was reviewed, and adjacent categories were collapsed to achieve a minimum cell size of n=50 in the exploratory set (see Supplementary Information for a list of response categories). The same transformations were applied to the confirmatory set.

#### 2.4.2 Exploratory analysis

Exploratory factor analysis (EFA) was conducted in the exploratory dataset using Mplus Version 8.4, and the R package MplusAutomation (16) was used to process the output. Given the ordinal nature of the data, we employed a categorical factor model with diagonally weighted least squares estimation with mean- and variance-adjusted chi-square statistics and standard errors, and geomin (oblique) rotation. Two approaches were used: standard EFA and bifactor EFA. The bifactor model specifies a general factor (always the first factor extracted), which in this case would approximate the “p factor” described by Caspi et al., and additional factors account for the remaining shared variation. The root mean square error of approximation (RMSEA), comparative fit index (CFI), and Tucker-Lewis index (TLI) were generated and interpreted as described below. We selected the best model from these candidates by first determining which solutions had relatively better fit, and then evaluating the clinical interpretability of each solution. Items were adopted onto a factor if they possessed a loading >0.30; in the standard EFA, cross-loading was allowed because it is theoretically consistent with the constructs measured by the DSM-XC.

#### 2.4.3 Confirmatory analyses

The confirmatory factor analysis (CFA) was conducted in the confirmatory dataset using lavaan version 0.6-7 (17) in R version 4.0.2. lavaan also employs diagonally weighted least squares estimation for ordinal data, and the Santorra-Bentler correction was applied to the test statistic. The robust fit indices (RMSEA, CFI, TLI) are reported and interpreted as described below.

#### 2.4.4 Invariance analyses

Measurement invariance analyses were conducted using semTools version 0.5-4 (18). We sequentially tested increasing levels of measurement invariance: configural, thresholds, metric, and scalar. Change in the relative (robust) fit indices RMSEA, CFI, and TLI were interpreted as described below. The measurement invariance of the selected solution across age (18-29 years, n = 1330; 40-59 years, n = 1370; 60+ years, n = 791) and sex (males, n=589; females, n = 2921) was evaluated using the full sample (combined testing and training, N = 3,533). Respondents missing age or sex data were excluded from the respective invariance analysis. We also evaluated the cross-sectional invariance of the solution across enrollment date, comparing baseline responses for participants whose study baseline occurred during the first month of the study (April 04 through May 04, 2020, n = 1380) to those who started during months 5 - 7 of the study (September 01 through October 31, 2020, n = 490).

#### 2.4.5 Interpretation of fit indices

While several sets of guidelines for the interpretation of fit indices exist (e.g. (19)), so do strong arguments against the use of cutoffs, especially when diagonally weighted least squares estimation was used (e.g. (20–24)). Without a universally agreed upon alternative, we chose to use the RMSEA, CFI, and TLI to guide our modelling decisions. Rather than classify these indices into “good” or “bad,” we evaluated each from a relative perspective. In other words, lower (scaled) chi-square to DF ratios, lower RMSEA, and higher TLI or CFI are preferred, regardless of their actual values.

The Mplus and R code files for all analyses are provided in Supplementary Information. For more information on how to interpret factor scores, please see (25).

## 3. Results

During the 7-month period of study recruitment, 3654 participants from all 50 U.S. states signed an electronic consent form and completed the study enrollment surveys. Of these participants, 3533 provided responses to all 22 DSM-XC items in our version of the survey and were included in this analysis. The mean age of this final sample was 46.5 +/-14.8 with a range from 18 to 87 years. The sample was majority female (83.2%) and white (90.2%). The majority of the sample (71.6%) endorsed a history of mental health counseling. Additionally, about half (53.2%) endorsed a history of medication for a mental health condition and 13.8% endorsed a past psychiatric hospitalization (Table 1).

**Table 1.** Demographics of sample

| Demographic Characteristics | Total Sample |
| --- | --- |
| Age (Range) | 46.5 ± 14.8 (18-87) |
| Sex |  |
| Male, *n* (%) | 589 (16.8%) |
| Female, *n* (%) | 2921 (83.2%) |
| Race |  |
| White, *n* (%) | 3150 (90.2%) |
| Black, *n* (%) | 104 (3.0%) |
| Asian, *n* (%) | 85 (2.4%) |
| Multiple, *n* (%) | 155 (4.4%) |
| Education |  |
| < college degree, *n* (%) | 395 (11.1%) |
| Associates, *n* (%) | 175 (5.0%) |
| Bachelors, *n* (%) | 1112 (31.6%) |
| Advanced, *n* (%) | 1840 (52.3%) |
| Self-Report Clinical History |  |
| Psych Hospitalization, *n* (%) | 487 (13.8%) |
| Psych Medication Use, *n* (%) | 1880 (53.2%) |
| Mental Health Counseling, *n* (%) | 2315 (71.6%) |

### Exploratory factor analysis

Based on clinical usage of the instrument and six eigenvalues greater than 1, we explored solutions with one through six factors in the training dataset (n = 1202). The fit indices for these solutions are shown in Table 2 and the standardized loadings for each solution are shown in Supplementary Tables 1 and 2. Based on fit indices and interpretability, we carried forward the six-factor standard EFA and the two-factor bifactor solutions.

**Table 2.** Fit indices

| | $\chi^2$ | df | CFI | TLI | RMSEA [95% CI] |
| --- | --- | --- | --- | --- | --- |
| *Exploratory Factor Analyses* |  |  |  |  |  |
| Standard Solutions (# factors) |  |  |  |  |  |
| 1 | 1408.03 | 209 | 0.952 | 0.947 | 0.069 [0.066, 0.070] |
| 2 | 931.02 | 188 | 0.97 | 0.964 | 0.057 [0.054, 0.061] |
| 3 | 535.89 | 168 | 0.985 | 0.98 | 0.043 [0.039, 0.047] |
| 4 | 372.34 | 149 | 0.991 | 0.986 | 0.035 [0.031, 0.040] |
| 5 | 244.78 | 131 | 0.995 | 0.992 | 0.027 [0.022, 0.032] |
| 6 | 186.79 | 114 | 0.997 | 0.994 | 0.023 [0.017, 0.029] |
| Bifactor Solutions (# factors) |  |  |  |  |  |
| 2 | 931.024 | 188 | 0.97 | 0.964 | 0.057 [0.054, 0.061] |
| 3 | 535.893 | 168 | 0.985 | 0.98 | 0.043 [0.039, 0.047] |
| 4 | 372.343 | 149 | 0.991 | 0.986 | 0.035 [0.031, 0.040] |
| 5 | 244.778 | 131 | 0.995 | 0.992 | 0.027 [0.022, 0.032] |
| 6 | 186.795 | 114 | 0.997 | 0.994 | 0.023 [0.017, 0.029] |
| *Confirmatory Factor Analyses* |  |  |  |  |  |
| Lace and Merz 2-Factor | 3690.06 | 208 | 0.982 | 0.98 | 0.062 [0.064, 0.066] |
| Proposed 6-Factor Solution | 1392.33 | 187 | 0.994 | 0.993 | 0.038 [0.036, 0.040] |
| Proposed Bifactor Solution | 1897.65 | 199 | 0.991 | 0.999 | 0.045 [0.044, 0.047] |

Notes: $\chi^2$ – Chi Square, df – Degrees of Freedom, CFI – Comparative Fit Index, TLI – Tucker Lewis Index, RMSEA – Root Mean Square Error of Approximation, SRMR - Standardized Root Mean Square Residual. Robust indices used for Confirmatory Factor Analyses.

### Confirmatory factor analysis

The results of the CFA indicated that both the six-factor and two-factor bifactor solutions were a good fit for the testing data (n = 2331; Table 2). Both models had good interpretability. We propose that the six factors represent the following constructs: mood, worry, activation, somatic, thoughts, and substance use (Figure 1). The bifactor model, in addition to the general psychopathology factor, represents internalizing and thought disorder constructs (Figure 2). The standardized loadings on each factor were moderate-to-high (Table 3). Finally, we evaluated the existing Lace and Merz (13) two-factor solution, and found that our data exhibited acceptable fit to this solution (Table 3).

**Table 3.** Standardized CFA loadings of selected solutions

| Item | Six-factor Solution |  |  |  |  |  | Bifactor Solution |  |  |
| --- | --- | --- | --- | --- | --- | --- | --- | --- | --- |
|  | Factor 1 | Factor 2 | Factor 3 | Factor 4 | Factor 5 | Factor 6 | General Factor | Factor 1 | Factor 2 |
| Little interest or pleasure in doing things? | 0.89 |  |  |  |  |  | 0.84 | 0.30 |  |
| Feeling down, depressed or hopeless? | 0.91 |  |  |  |  |  | 0.86 | 0.29 |  |
| Feeling more irritated, grouchy, or angry than usual? | 0.75 |  |  |  |  |  | 0.72 |  |  |
| Not knowing who you really are or what you want out of life? | 0.67 |  |  |  | 0.37 |  | 0.72 |  |  |
| Not feeling close to other people or enjoying relationships? | 0.71 |  |  |  | 0.31 |  | 0.75 |  |  |
| Feeling detached from yourself, your body, your surroundings? | 0.69 |  |  |  | 0.37 |  | 0.74 |  |  |
| Feeling nervous, anxious, worried or on edge? |  | 0.91 |  |  |  |  | 0.76 |  | 0.48 |
| Feeling panic or being frightened? |  | 0.90 |  |  |  |  | 0.74 |  | 0.59 |
| Avoiding situations that make you anxious? |  | 0.78 |  |  |  |  | 0.67 |  | 0.32 |
| Feeling driven to perform certain behaviors over and over? |  | 0.60 |  |  | 0.48 |  | 0.66 | -0.24 |  |
| Unpleasant thoughts, urges or images repeatedly enter your mind? |  | 0.71 |  |  | 0.52 |  | 0.75 |  |  |
| Starting more projects or doing more risky things than usual? |  |  | 0.57 |  |  |  | 0.45 | -0.59 |  |
| Sleeping less than usual, but still have a lot of energy? |  |  | 0.43 |  |  |  | 0.33 | -0.49 |  |
| Problems with sleep that affected your sleep quality over all? |  |  | 0.75 |  |  |  | 0.57 |  |  |
| Problems with memory or with location? |  |  |  | 0.74 |  |  | 0.67 |  |  |
| Unexplained aches and pains? |  |  |  | 0.67 |  |  | 0.61 |  |  |
| Feeling that your illnesses are not being taken seriously? |  |  |  | 0.78 |  |  | 0.70 |  |  |
| Hearing things that other people couldn't hear, such as voices? |  |  |  | 0.56 | 0.35 |  | 0.58 | -0.39 |  |
| Feeling that someone could hear your thoughts? |  |  |  | 0.51 | 0.36 |  | 0.55 | -0.41 |  |
| Drinking at least 4 drinks of alcohol in a single day? |  |  |  |  |  | 0.32 | 0.17 |  |  |
| Smoking cigarettes, a cigar, pipe, using snug or chewing tobacco? |  |  |  |  |  | 0.68 | 0.39 |  |  |
| Using medications without a doctor's prescription? |  |  |  |  |  | 0.74 | 0.42 |  |  |

Note: Some items have been shortened. Items are standardized loadings from confirmatory factor analysis.

**Figure 1.**
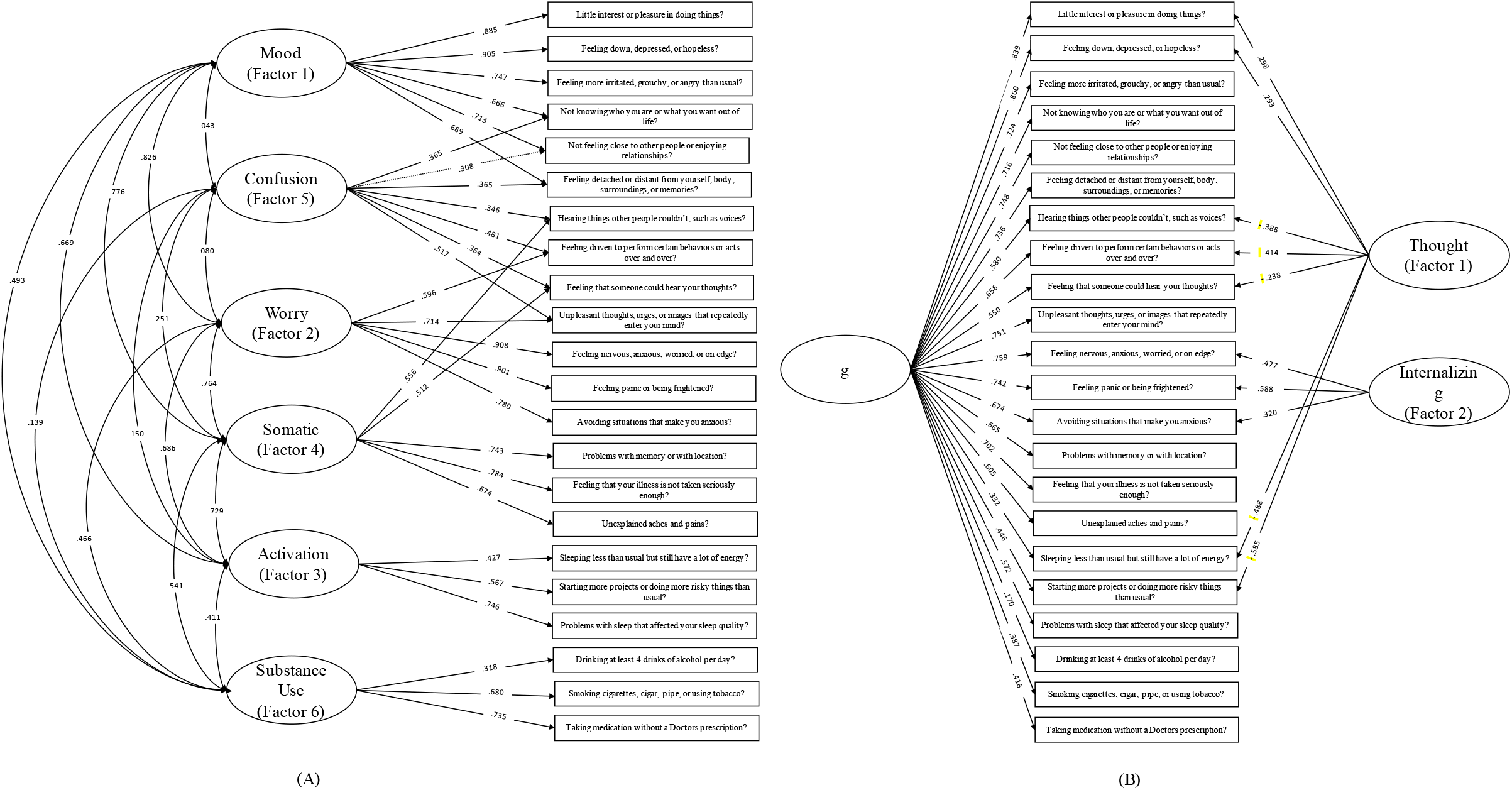
(A) Simplified path diagram for six-factor solution. Rectangles represent items (names have been shortened). Circles represent factors. Standardized loadings are shown. (Simplified path diagram for bifactor solution. Solid rectangles represent items (names have been shortened). Circles represent factors. Standardized factor loadings are shown; negative loadings have been highlighted by a dotted rectangle.

### Measurement invariance

Configural, threshold, metric, and scalar measurement invariance of both the six-factor and bifactor solutions were evaluated sequentially across age (n = 3491), sex (n = 3510), and enrollment date (n = 1870). Owing to the smaller sample size for the enrollment date analyses, too few non-zero responses to the items “Hearing things…” and “…hear your thoughts” meant that they had to be excluded from the solution. For both models, full measurement invariance was supported across age, sex, and enrollment date (Table 4).

**Table 4.** Results of measurement invariance analyses

| Grouping | Model | Six-Factor Solution |  |  |  | Bifactor Solution |  |  |  |
| --- | --- | --- | --- | --- | --- | --- | --- | --- | --- |
|  |  | df | CFI | TLI | RMSEA | df | CFI | TLI | RMSEA |
| Age | Configural | 561 | 0.980 | 0.975 | 0.042 | 597 | 0.971 | 0.966 | 0.049 |
|  | Threshold | 605 | 0.979 | 0.976 | 0.042 | 641 | 0.970 | 0.968 | 0.048 |
|  | Metric | 651 | 0.982 | 0.981 | 0.038 | 699 | 0.978 | 0.979 | 0.039 |
|  | Scalar | 671 | 0.981 | 0.981 | 0.038 | 728 | 0.976 | 0.978 | 0.040 |
| Sex | Configural | 374 | 0.987 | 0.983 | 0.038 | 398 | 0.978 | 0.975 | 0.046 |
|  | Threshold | 399 | 0.986 | 0.984 | 0.037 | 423 | 0.978 | 0.976 | 0.045 |
|  | Metric | 422 | 0.987 | 0.986 | 0.034 | 452 | 0.980 | 0.980 | 0.041 |
|  | Scalar | 430 | 0.987 | 0.986 | 0.034 | 466 | 0.982 | 0.982 | 0.039 |
| Enrollment Date | Configural | 300 | 0.984 | 0.980 | 0.043 | 324 | 0.974 | 0.969 | 0.053 |
|  | Threshold | 316 | 0.984 | 0.981 | 0.042 | 340 | 0.973 | 0.970 | 0.052 |
|  | Metric | 335 | 0.985 | 0.983 | 0.039 | 365 | 0.978 | 0.977 | 0.046 |
|  | Scalar | 340 | 0.986 | 0.984 | 0.038 | 376 | 0.979 | 0.978 | 0.045 |

Note: CFI = comparative fit index (higher is better); TLI = Tucker-Lewis fit index (higher is better); RMSEA = root mean square error of approximation (lower is better)

## 4. Discussion

In this secondary analysis, we explored the factor structure of the DSM-XC using a large data set of individuals with varying degrees of psychological health. While the previously published solution of internalizing and externalizing factors (13) had a mediocre fit to our data, we evaluated the evidence for the factorial validity and measurement invariance of two other multidimensional solutions. Given the lack of literature on the DSM-XC and its latent dimensionality, we believe these findings will help future researchers score and interpret DSM-XC responses.

While the existing 13-domain structure of the DSM-XC would have been an obvious candidate for evaluation, such an analysis was not possible because several domains include only one or two items. As with any factor analysis, alternative structures may provide similar fit to the data. However, both candidate solutions we propose herein suggest that these items are better conceptualized as fewer, broader constructs. Both the six-factor and bifactor solutions exhibited strong factorial validity, demonstrated through a confirmatory analysis in a withheld subset of our data and a series of measurement invariance analyses.

We argue that the six-factor solution holds considerable face validity and appears to be clinically coherent based on clinician consensus review of symptom item clusters. The items on each factor suggest that they represent the following constructs (respectively): mood, worry, activation, somatic, thoughts, and substance use (Figure 1). In most cases, items which shared a domain on the DSM-XC also shared a factor in our proposed solution (e.g., the two depression items load onto factor 1, three anxiety items load onto factor 2).

The results of our bifactor analysis are theoretically consistent with findings from Caspi and colleagues (15). The general psychopathology factor is a satisfying solution given the known overlap of symptoms across psychiatric disorders and their comorbidity. While the general psychopathology factor explained most of the variance, two residual factors were identified. The first included the three items from the DSM-XC anxiety domain, which we termed “internalizing.” The second included items that index psychosis, mania and depression symptoms which we labeled “thought disorder.” Both labels were chosen to be consistent with the Caspi model.

Our findings are particularly relevant for use of the DSM-XC since it has been promoted as a dimensional measure of psychopathology (5). Our findings first confirm that the DSM-XC is a multidimensional instrument. While researchers may decide to use the DSM-XC as an indicator of overall symptom burden and simply tally the total score of all items, all available factor analyses of the instrument, including ours, indicate that the scale is not unidimensional. Even considering the general factor in the multidimensional bifactor solution, we note that some items load much more strongly on the general factor than others, a fact obscured by the simple sum total of scores. Further, the use of factor scores, rather than subscale totals, enable researchers to measure with more precision the dimensional mental health constructs reflected by the scale. Second, our findings provide a scoring solution for future researchers who wish to measure distinct mental health constructs. While thirteen clinically derived domains are provided, our findings suggest that there are fewer, broader constructs, and fitting with our conceptualization of mental health, that many items are not unique to a particular construct. This solution therefore accounts for cross-cutting symptoms that relate to multiple constructs of interest. Future research should evaluate the structures proposed in this paper to confirm its consistency across research populations and settings. An additional important future direction is the evaluation of the validity of the proposed solutions with convergent measures; such an analysis was outside the scope of this secondary analysis.

Several limitations stem from the fact that this study was conducted online during a global pandemic. First, given constraints on recruitment methods during the COVID-19 pandemic, we enrolled a convenience sample with demographic characteristics not representative of the general population (i.e., 90% white, 83% female, 52% advanced degree). It is also important to note that our population was likely subject to sampling bias due to the study’s focus on mental health: over 50% of our sample endorsed a history of psychiatric medication use, which is substantially higher than rates of use in the general population (26). Similarly, factor scores are likely elevated due to the COVID-19 pandemic (27,28). Finally, this study used a modified version of the DSM-XC, as we chose to omit the self-harm item.

Despite these limitations, this study contributes to the growing body of literature on the DSM-XC and can help with the interpretation of scores for future research. The measure has been recommended for clinical research by the NIMH Extramural Research Program (7) and for further evaluation by the APA (29). Continued evaluation of its psychometric properties is supported by a need to better identify mental health in the general population; while many domain-specific mental health surveys have been validated, the DSM-XC captures symptoms that cross-cut psychiatric diagnostic categories which accounts for the dimensionality and frequent co-occurrence of mental illnesses.

## Supporting information

Supplemental file loadings and syntax

## Data Availability

A NIMH Data Sharing form has been completed and submitted in accordance with the NIH Intramural Research policy on broad sharing of NIH research data.These data will be made available after publication of study findings.

## Acknowledgements

The research was funded by the Intramural Research Program of the National Institute of Mental Health (ZIAMH002922). The sponsor had no role in study design; in the collection, analysis, and interpretation of data; in the writing of the report; and in the decision to submit the article for publication.

## Notes

### Competing Interest Statement

The authors have declared no competing interest.

### Clinical Trial

NCT04339790

### Clinical Protocols

https://nimhcovidstudy.ctss.nih.gov/

### Summary of Updates

We used a factor analytic framework to identify an bifactor candidate solution in addition to the six-factor solution we reported in the previous version of the manuscript.

## References

1. Substance Abuse and Mental Health Services Administration. Key Substance Use and Mental Health Indicators in the United States: Results from the 2019 National Survey on Drug Use and Health | SAMHSA Publications and Digital Products [Internet]. 2020 [cited 2021 Feb 24]. Available from: https://store.samhsa.gov/product/key-substance-use-and-mental-health-indicators-in-the-united-states-results-from-the-2019-national-survey-on-Drug-Use-and-Health/PEP20-07-01-001

2. Kroenke K, Spitzer RL, Williams JBW. The PHQ-9. J Gen Intern Med. 2001 Sep;16(9):606–13.

3. Jones KD. Dimensional and Cross-Cutting Assessment in the DSM-5. J Couns Dev. 2012 Oct;90(4):481–7.

4. Narrow WE, Clarke DE, Kuramoto SJ, Kraemer HC, Kupfer DJ, Greiner L, et al. DSM-5 Field Trials in the United States and Canada, Part III: Development and Reliability Testing of a Cross-Cutting Symptom Assessment for DSM-5. Am J Psychiatry. 2013 Jan 1;170(1):71–82.

5. Clarke DE, Kuhl EA. DSM-5 cross-cutting symptom measures: a step towards the future of psychiatric care? World Psychiatry. 2014 Oct;13(3):314–6.

6. Mościcki EK, Clarke DE, Kuramoto SJ, Kraemer HC, Narrow WE, Kupfer DJ, et al. Testing DSM-5 in Routine Clinical Practice Settings: Feasibility and Clinical Utility. Psychiatr Serv. 2013 Oct 1;64(10):952–60.

7. NOT-MH-15-009: Notice Announcing Data Harmonization for NIMH Human Subjects Research via the PhenX Toolkit [Internet]. 2021 [cited 2021 Feb 24]. Available from: https://grants.nih.gov/grants/guide/notice-files/NOT-MH-15-009.html

8. Mahoney MR, Farmer C, Sinclair S, Sung S, Dehaut K, Chung JY. Utilization of the DSM-5 Self-Rated Level 1 Cross-Cutting Symptom Measure-Adult to Screen Healthy Volunteers for Research Studies. Psychiatry Res. 2020 Apr;286:112822.

9. Bastiaens L, Galus J. The DSM-5 Self-Rated Level 1 Cross-Cutting Symptom Measure as a Screening Tool. Psychiatr Q. 2018 Mar;89(1):111–5.

10. Bravo A, Villarose-Hurlocker M, Pearson M, Protective Strategies Study Team. College Student Mental Health: An Evaluation of the DSM–5 Self-Rated Level 1 Cross-Cutting Symptom Measure. Psychol Assesment. 2018;30(10):1382–9.

11. Çökmüş F, Balıkçı K, Aydemir Ö, Grubu D. Reliability and validity of Turkish Form of DSM-5 Self-Rated Level 1 Cross-Cutting Symptom Scale-Adult Version. Anatol J Psychiatry. 2017;18(2):5.

12. Ishfaq N, Kamal A. Mental health and imprisonment: Measuring cross-cutting symptoms among convicts in Punjab, Pakistan. Asian J Psychiatry. 2019 Aug 1;44:127–32.

13. Lace JW, Merz ZC. DSM-5 Level 1 cross-cutting measure in an online sample: evaluating its latent dimensionality and utility detecting nonspecific psychological distress. Psychiatry Res. 2020 Dec;294:113529.

14. Costello A, Osborne J. Best practices in exploratory factor analysis: four recommendations for getting the most from your analysis. Pract Assess Res Eval [Internet]. 2019 Nov 23;10(1). Available from: https://scholarworks.umass.edu/pare/vol10/iss1/7

15. Caspi A, Houts RM, Belsky DW, Goldman-Mellor SJ, Harrington H, Israel S, et al. The p Factor: One General Psychopathology Factor in the Structure of Psychiatric Disorders? Clin Psychol Sci. 2014 Mar;2(2):119–37.

16. Hallquist MN, Wiley JF. MplusAutomationLJ: An R Package for Facilitating Large-Scale Latent Variable Analyses in M plus. Struct Equ Model Multidiscip J. 2018 Jul 4;25(4):621–38.

17. Rosseel Y. lavaan: An R Package for Structural Equation Modeling. J Stat Softw. 2012 May 24;48(1):1–36.

18. Jorgensen TD, Pornprasertmanit S, Schoemann AM, Rosseel Y, Miller P, Quick C, et al. semTools: Useful Tools for Structural Equation Modeling [Internet]. 2021 [cited 2021 Mar 28]. Available from: https://CRAN.R-project.org/package=semTools

19. Hu L, Bentler PM. Cutoff criteria for fit indexes in covariance structure analysis: Conventional criteria versus new alternatives. Struct Equ Model Multidiscip J. 1999 Jan 1;6(1):1–55.

20. Barrett P. Structural equation modelling: Adjudging model fit. Personal Individ Differ. 2007 May;42(5):815–24.

21. Cook KF, Kallen MA, Amtmann D. Having a fit: impact of number of items and distribution of data on traditional criteria for assessing IRT’s unidimensionality assumption. Qual Life Res Int J Qual Life Asp Treat Care Rehabil. 2009 May;18(4):447–60.

22. Marsh HW, Wen Z, Hau K-T. Structural Equation Models of Latent Interactions: Evaluation of Alternative Estimation Strategies and Indicator Construction. Psychol Methods. 2004;9(3):275–300.

23. McIntosh CN. Rethinking fit assessment in structural equation modelling: A commentary and elaboration on Barrett (2007). Personal Individ Differ. 2007 May;42(5):859–67.

24. Xia Y, Yang Y. RMSEA, CFI, and TLI in structural equation modeling with ordered categorical data: The story they tell depends on the estimation methods. Behav Res Methods. 2019 Feb 1;51(1):409–28.

25. DiStefano C, Zhu M, Mîndrilã D. Understanding and Using Factor Scores: Considerations for the Applied Researcher. Pract Assess Res Eval [Internet]. 2019 Nov 23;14(1). Available from: https://scholarworks.umass.edu/pare/vol14/iss1/20

26. Terlizzi E, Zablotsky B. Mental Health Treatment Among Adults: United States, 2019. US Department of Health and Human Services, Centers for Disease Control and Prevention; 2020 Sep p. https://www.cdc.gov/nchs/products/databriefs/db380.htm. Report No.: 380.

27. Panchal N, Kamal R, 2021. The Implications of COVID-19 for Mental Health and Substance Use [Internet]. KFF. 2021 [cited 2021 Feb 24]. Available from: https://www.kff.org/coronavirus-covid-19/issue-brief/the-implications-of-covid-19-for-mental-health-and-substance-use/

28. Xiong J, Lipsitz O, Nasri F, Lui LMW, Gill H, Phan L, et al. Impact of COVID-19 pandemic on mental health in the general population: A systematic review. J Affect Disord. 2020 Dec 1;277:55–64.

29. American Psychiatric Association. Online Assessment Measures [Internet]. 2019 [cited 2021 Feb 24]. Available from: https://www.psychiatry.org/psychiatrists/practice/dsm/educational-resources/assessment-measures

