## Supplemental file loadings and syntax for "Examining the Factor Structure of the DSM-5 Level 1 Cross-Cutting Symptom Measure"

Contents

Table S1: Standardized loadings for exploratory factor analysis solutions

Table S2: Standardized loadings for exploratory bifactor analysis solutions

Model syntax 1: Exploratory factor analysis (Mplus)

Model syntax 2: Exploratory bifactor analysis (Mplus)

Model syntax 3: Confirmatory factor analysis (R)

Model syntax 4: Measurement invariance (R)

**Table S1: Standardized loadings for exploratory factor analysis solutions.** Note that some item names have been shortened.

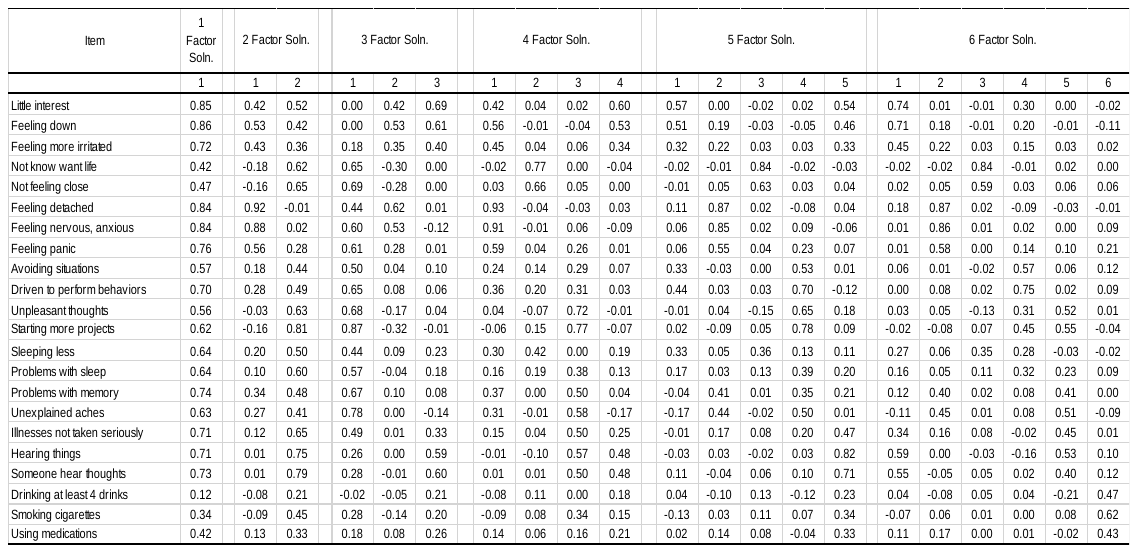

**Table S2: Standardized loadings for exploratory bifactor analysis solutions.** Note that some item names have been shortened.

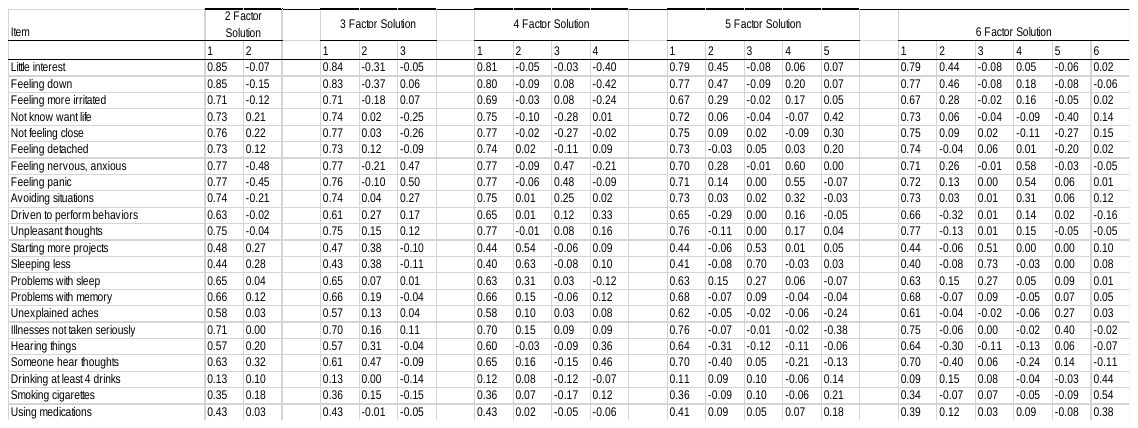

**Model syntax 1: Exploratory factor analysis (Mplus)**

title: DSM XC EFA

data: file is TRAIN.TXT;

variable:

names are ID

LITTLE_INTEREST

DOWN_HOPELESS

MORE_IRRITATED

SLEEPNG_LESS

DOING_MORE

NERVOUS_ANXIOUS

PANIC

AVOIDNG_ANXIOUS

UNEXPLAIND_PAIN

ILLNESS_NOT_TAKEN_SERIOUSLY

HEAR_THINGS

HEAR_THOUGHTS

PROB_SLEEP

PROB_MEMORY

UNPLEASANT_THOUGHTS

PERFORM_REPEATNG_ACTS

DISTANT_SELF

NOT_LIFE_WANT

NOT_FEELNG_CLOSE

MORE_4_DRINK_PER_DAY

TOBACCO

SELF_MEDICATED;

categorical are LITTLE_INTEREST

DOWN_HOPELESS

MORE_IRRITATED

SLEEPNG_LESS

DOING_MORE

NERVOUS_ANXIOUS

PANIC

AVOIDNG_ANXIOUS

UNEXPLAIND_PAIN

ILLNESS_NOT_TAKEN_SERIOUSLY

HEAR_THINGS

HEAR_THOUGHTS

PROB_SLEEP

PROB_MEMORY

UNPLEASANT_THOUGHTS

PERFORM_REPEATNG_ACTS

DISTANT_SELF

NOT_LIFE_WANT

NOT_FEELNG_CLOSE

MORE_4_DRINK_PER_DAY

TOBACCO

SELF_MEDICATED;

usevariables

LITTLE_INTEREST

DOWN_HOPELESS

MORE_IRRITATED

SLEEPNG_LESS

DOING_MORE

NERVOUS_ANXIOUS

PANIC

AVOIDNG_ANXIOUS

UNEXPLAIND_PAIN

ILLNESS_NOT_TAKEN_SERIOUSLY

HEAR_THINGS

HEAR_THOUGHTS

PROB_SLEEP

PROB_MEMORY

UNPLEASANT_THOUGHTS

PERFORM_REPEATNG_ACTS

DISTANT_SELF

NOT_LIFE_WANT

NOT_FEELNG_CLOSE

MORE_4_DRINK_PER_DAY

TOBACCO

SELF_MEDICATED;

analysis: type = efa 1 6;

**Model syntax 2: Exploratory bifactor analysis (Mplus)**

title: DSM XC bifactor efa

data: file is TRAIN.TXT;

variable:

names are ID

LITTLE_INTEREST

DOWN_HOPELESS

MORE_IRRITATED

SLEEPNG_LESS

DOING_MORE

NERVOUS_ANXIOUS

PANIC

AVOIDNG_ANXIOUS

UNEXPLAIND_PAIN

ILLNESS_NOT_TAKEN_SERIOUSLY

HEAR_THINGS

HEAR_THOUGHTS

PROB_SLEEP

PROB_MEMORY

UNPLEASANT_THOUGHTS

PERFORM_REPEATNG_ACTS

DISTANT_SELF

NOT_LIFE_WANT

NOT_FEELNG_CLOSE

MORE_4_DRINK_PER_DAY

TOBACCO

SELF_MEDICATED;

categorical are LITTLE_INTEREST

DOWN_HOPELESS

MORE_IRRITATED

SLEEPNG_LESS

DOING_MORE

NERVOUS_ANXIOUS

PANIC

AVOIDNG_ANXIOUS

UNEXPLAIND_PAIN

ILLNESS_NOT_TAKEN_SERIOUSLY

HEAR_THINGS

HEAR_THOUGHTS

PROB_SLEEP

PROB_MEMORY

UNPLEASANT_THOUGHTS

PERFORM_REPEATNG_ACTS

DISTANT_SELF

NOT_LIFE_WANT

NOT_FEELNG_CLOSE

MORE_4_DRINK_PER_DAY

TOBACCO

SELF_MEDICATED;

usevariables

LITTLE_INTEREST

DOWN_HOPELESS

MORE_IRRITATED

SLEEPNG_LESS

DOING_MORE

NERVOUS_ANXIOUS

PANIC

AVOIDNG_ANXIOUS

UNEXPLAIND_PAIN

ILLNESS_NOT_TAKEN_SERIOUSLY

HEAR_THINGS

HEAR_THOUGHTS

PROB_SLEEP

PROB_MEMORY

UNPLEASANT_THOUGHTS

PERFORM_REPEATNG_ACTS

DISTANT_SELF

NOT_LIFE_WANT

NOT_FEELNG_CLOSE

MORE_4_DRINK_PER_DAY

TOBACCO

SELF_MEDICATED;

ANALYSIS: TYPE = EFA 2 6;

ROTATION = BI-GEOMIN;

**Model syntax 3: Confirmatory factor analysis (R)**

library(tidyverse)

library(lavaan)

\# Lace and Merz model

factor.LM <- 'internalizing =~ LITTLE_INTEREST + DOWN_HOPELESS + MORE_IRRITATED + SLEEPNG_LESS + NERVOUS_ANXIOUS + PANIC + AVOIDNG_ANXIOUS + UNEXPLAIND_PAIN + ILLNESS_NOT_TAKEN_SERIOUSLY + PROBLEMS_SLEEP + UNPLEASANT_THOUGHTS + NOT_LIFE_WANT + NOT_FEELNG_CLOSE

externalizing =~ DOING_MORE + HEAR_THOUGHTS + HEARING_THINGS + PROBLEMS_MEMORY + PERFORM_REPEATNG_ACTS + DISTANT_SELF + MORE_4_DRINK_PER_DAY + TOBACCO + SELF_MEDICATED'

\# NIH six-factor model

nih6cross <- 'Factor1 =~ LITTLE_INTEREST + DOWN_HOPELESS + MORE_IRRITATED + NOT_LIFE_WANT + NOT_FEELNG_CLOSE + DISTANT_SELF

Factor2 =~ NERVOUS_ANXIOUS + PANIC + AVOIDNG_ANXIOUS + PERFORM_REPEATNG_ACTS + UNPLEASANT_THOUGHTS

Factor3 =~ DOING_MORE + SLEEPNG_LESS + PROBLEMS_SLEEP

Factor4 =~ PROBLEMS_MEMORY + UNEXPLAIND_PAIN + ILLNESS_NOT_TAKEN_SERIOUSLY + HEARING_THINGS + HEAR_THOUGHTS

Factor5 =~ HEARING_THINGS + HEAR_THOUGHTS + PERFORM_REPEATNG_ACTS + UNPLEASANT_THOUGHTS + DISTANT_SELF + NOT_LIFE_WANT + NOT_FEELNG_CLOSE

Factor6 =~ MORE_4_DRINK_PER_DAY + TOBACCO + SELF_MEDICATED'

\# NIH bifactor model

bifactor <- "g =~ DOWN_HOPELESS + LITTLE_INTEREST + DOWN_HOPELESS + MORE_IRRITATED + SLEEPNG_LESS + DOING_MORE + NERVOUS_ANXIOUS + PANIC + AVOIDNG_ANXIOUS + UNEXPLAIND_PAIN + ILLNESS_NOT_TAKEN_SERIOUSLY + HEARING_THINGS + HEAR_THOUGHTS + PROBLEMS_SLEEP + PROBLEMS_MEMORY + UNPLEASANT_THOUGHTS + PERFORM_REPEATNG_ACTS + DISTANT_SELF + NOT_LIFE_WANT + NOT_FEELNG_CLOSE + MORE_4_DRINK_PER_DAY + TOBACCO + SELF_MEDICATED

Factor1 =~ LITTLE_INTEREST + DOWN_HOPELESS + SLEEPNG_LESS + DOING_MORE + HEARING_THINGS + HEAR_THOUGHTS + PERFORM_REPEATNG_ACTS

Factor2 =~ NERVOUS_ANXIOUS + PANIC + AVOIDNG_ANXIOUS

g ~~ 0*Factor1

g ~~ 0*Factor2

Factor1 ~~ 0*Factor2"

#CFA for Lace & Merz model

fit.LM <- cfa(model=factor.LM, data = DSM.data, test = "Satorra-Bentler")

summary(fit.LM)

#CFA for NIH 6 factor

fit.6.cross <- cfa(model=nih6cross, data = DSM.data, test = "Satorra-Bentler")

summary(fit.6.cross)

#CFA for NIH bifactor model

fit.g <- cfa(bifactor, data = DSM.data, test = "Satorra-Bentler")

summary(fit.g)

**Model syntax 4: Measurement invariance (R)**

library(tidyverse)

library(lavaan)

library(semTools)

\# selected 6-factor solution

nih6cross <- '

Factor1 =~ LITTLE_INTEREST + DOWN_HOPELESS + MORE_IRRITATED + NOT_LIFE_WANT + NOT_FEELNG_CLOSE + DISTANT_SELF

Factor2 =~ NERVOUS_ANXIOUS + PANIC + AVOIDNG_ANXIOUS + PERFORM_REPEATNG_ACTS + UNPLEASANT_THOUGHTS

Factor3 =~ DOING_MORE + SLEEPNG_LESS + PROBLEMS_SLEEP

Factor4 =~ PROBLEMS_MEMORY + UNEXPLAIND_PAIN + ILLNESS_NOT_TAKEN_SERIOUSLY + HEARING_THINGS + HEAR_THOUGHTS

Factor5 =~ HEARING_THINGS + HEAR_THOUGHTS + PERFORM_REPEATNG_ACTS + UNPLEASANT_THOUGHTS + DISTANT_SELF + NOT_LIFE_WANT + NOT_FEELNG_CLOSE

Factor6 =~ MORE_4_DRINK_PER_DAY + TOBACCO + SELF_MEDICATED'

\# selected 6-factor solution excluding hearing things and hearing thoughts which have zero variance in enrollment date set

nih6cross.sub <- '

Factor1 =~ LITTLE_INTEREST + DOWN_HOPELESS + MORE_IRRITATED + NOT_LIFE_WANT + NOT_FEELNG_CLOSE + DISTANT_SELF

Factor2 =~ NERVOUS_ANXIOUS + PANIC + AVOIDNG_ANXIOUS + PERFORM_REPEATNG_ACTS + UNPLEASANT_THOUGHTS

Factor3 =~ DOING_MORE + SLEEPNG_LESS + PROBLEMS_SLEEP

Factor4 =~ PROBLEMS_MEMORY + UNEXPLAIND_PAIN + ILLNESS_NOT_TAKEN_SERIOUSLY

Factor5 =~ PERFORM_REPEATNG_ACTS + UNPLEASANT_THOUGHTS + DISTANT_SELF + NOT_LIFE_WANT + NOT_FEELNG_CLOSE

Factor6 =~ MORE_4_DRINK_PER_DAY + TOBACCO + SELF_MEDICATED'

\# selected bifactor solution

\# exchanged order of first two items on g so LITTLE_INTEREST not first item on both factors

bifactor <- "

g =~ DOWN_HOPELESS + LITTLE_INTEREST + DOWN_HOPELESS + MORE_IRRITATED + SLEEPNG_LESS + DOING_MORE + NERVOUS_ANXIOUS + PANIC + AVOIDNG_ANXIOUS + UNEXPLAIND_PAIN + ILLNESS_NOT_TAKEN_SERIOUSLY + HEARING_THINGS + HEAR_THOUGHTS + PROBLEMS_SLEEP + PROBLEMS_MEMORY + UNPLEASANT_THOUGHTS + PERFORM_REPEATNG_ACTS + DISTANT_SELF + NOT_LIFE_WANT + NOT_FEELNG_CLOSE + MORE_4_DRINK_PER_DAY + TOBACCO + SELF_MEDICATED

Factor1 =~ LITTLE_INTEREST + DOWN_HOPELESS + SLEEPNG_LESS + DOING_MORE + HEARING_THINGS + HEAR_THOUGHTS + PERFORM_REPEATNG_ACTS

Factor2 =~ NERVOUS_ANXIOUS + PANIC + AVOIDNG_ANXIOUS

g ~~ 0*Factor1

g ~~ 0*Factor2

Factor1 ~~ 0*Factor2"

\# selectd bifactor excl. hearing thoughts/hearing things which have zero variance in enrollment date set

bifactor.sub <- "

g =~ DOWN_HOPELESS + LITTLE_INTEREST + DOWN_HOPELESS + MORE_IRRITATED + SLEEPNG_LESS + DOING_MORE + NERVOUS_ANXIOUS + PANIC + AVOIDNG_ANXIOUS + UNEXPLAIND_PAIN + ILLNESS_NOT_TAKEN_SERIOUSLY + PROBLEMS_SLEEP + PROBLEMS_MEMORY + UNPLEASANT_THOUGHTS + PERFORM_REPEATNG_ACTS + DISTANT_SELF + NOT_LIFE_WANT + NOT_FEELNG_CLOSE + MORE_4_DRINK_PER_DAY + TOBACCO + SELF_MEDICATED

Factor1 =~ LITTLE_INTEREST + DOWN_HOPELESS + SLEEPNG_LESS + DOING_MORE + PERFORM_REPEATNG_ACTS

Factor2 =~ NERVOUS_ANXIOUS + PANIC + AVOIDNG_ANXIOUS

g ~~ 0*Factor1

g ~~ 0*Factor2

Factor1 ~~ 0*Factor2"

#mod.cat is the model nih6cross.sub, nih6cross, bifactor

#df.cat is the dataset long_bl (not actually a longitudinal dataset), recode_DSMage, recodeDSMsex

#group is the grouping variable in quotes "sex","agegroup","monthgrp"

createsyntax <- function(mod.cat, df.cat, group){

syntax.config <- measEq.syntax(configural.model = mod.cat,

data = df.cat,

parameterization = "theta",

ID.fac = "std.lv",

ID.cat = "Wu.Estabrook.2016",

group = group)

syntax.thresh <- measEq.syntax(configural.model = mod.cat,

data = df.cat,

parameterization = "theta",

ID.fac = "std.lv",

ID.cat = "Wu.Estabrook.2016",

group = group,

group.equal = "thresholds")

syntax.metric <- measEq.syntax(configural.model = mod.cat,

data = df.cat,

parameterization = "theta",

ID.fac = "std.lv",

ID.cat = "Wu.Estabrook.2016",

group = group,

group.equal = c("thresholds","loadings"))

syntax.scalar <- measEq.syntax(configural.model = mod.cat,

data = df.cat,

parameterization = "theta",

ID.fac = "std.lv",

ID.cat = "Wu.Estabrook.2016",

group = group,

group.equal = c("thresholds","loadings","intercepts"))

out = list(syntax.config,syntax.thresh,syntax.metric,syntax.scalar)

return(out)

} # syntax.scalar will throw error with crossloadings, see next chunk

syntax.age.cross = createsyntax(nih6cross, recode_DSMage, "agegroup")

syntax.age.bifactor = createsyntax(bifactor, recode_DSMage, "agegroup")

syntax.sex.cross = createsyntax(nih6cross, recode_DSMsex, "sex")

syntax.sex.bifactor = createsyntax(bifactor, recode_DSMsex, "sex")

syntax.time.cross = createsyntax(nih6cross.sub, long_bl, "monthgrp") #omit the hearing items

syntax.time.bifactor = createsyntax(bifactor.sub, long_bl, "monthgrp") #omit the hearing items

#manually free latent means for scalar model (necessary because of cross loading)

syntax.age.cross[[4]] <- update(syntax.age.cross[[4]],

change.syntax = "Factor1 ~ c(NA, NA, NA)*1 + c(alpha.1.g1, alpha.1.g2, alpha.1.g3)*1

Factor2 ~ c(NA, NA, NA)*1 + c(alpha.2.g1, alpha.2.g2, alpha.2.g3)*1

Factor3 ~ c(NA, NA, NA)*1 + c(alpha.3.g1, alpha.3.g2, alpha.3.g3)*1

Factor4 ~ c(NA, NA, NA)*1 + c(alpha.4.g1, alpha.4.g2, alpha.4.g3)*1

Factor5 ~ c(NA, NA, NA)*1 + c(alpha.5.g1, alpha.5.g2, alpha.5.g3)*1

Factor6 ~ c(NA, NA, NA)*1 + c(alpha.6.g1, alpha.6.g2, alpha.6.g3)*1")

syntax.age.bifactor[[4]] <- update(syntax.age.bifactor[[4]],

change.syntax = "g ~ c(NA, NA, NA)*1 + c(alpha.1.g1, alpha.1.g2, alpha.1.g3)*1

Factor1 ~ c(NA, NA, NA)*1 + c(alpha.2.g1, alpha.2.g2, alpha.2.g3)*1

Factor2 ~ c(NA, NA, NA)*1 + c(alpha.3.g1, alpha.3.g2, alpha.3.g3)*1")

syntax.sex.cross[[4]] <- update(syntax.sex.cross[[4]],

change.syntax = "Factor1 ~ c(NA, NA)*1 + c(alpha.1.g1, alpha.1.g2)*1

Factor2 ~ c(NA, NA)*1 + c(alpha.2.g1, alpha.2.g2)*1

Factor3 ~ c(NA, NA)*1 + c(alpha.3.g1, alpha.3.g2)*1

Factor4 ~ c(NA, NA)*1 + c(alpha.4.g1, alpha.4.g2)*1

Factor5 ~ c(NA, NA)*1 + c(alpha.5.g1, alpha.5.g2)*1

Factor6 ~ c(NA, NA)*1 + c(alpha.6.g1, alpha.6.g2)*1")

syntax.sex.bifactor[[4]] <- update(syntax.sex.bifactor[[4]],

change.syntax = "g ~ c(NA, NA)*1 + c(alpha.1.g1, alpha.1.g2)*1

Factor1 ~ c(NA, NA)*1 + c(alpha.2.g1, alpha.2.g2)*1

Factor2 ~ c(NA, NA)*1 + c(alpha.3.g1, alpha.3.g2)*1")

syntax.time.cross[[4]] <- update(syntax.time.cross[[4]],

change.syntax = "Factor1 ~ c(NA, NA)*1 + c(alpha.1.g1, alpha.1.g2)*1

Factor2 ~ c(NA, NA)*1 + c(alpha.2.g1, alpha.2.g2)*1

Factor3 ~ c(NA, NA)*1 + c(alpha.3.g1, alpha.3.g2)*1

Factor4 ~ c(NA, NA)*1 + c(alpha.4.g1, alpha.4.g2)*1

Factor5 ~ c(NA, NA)*1 + c(alpha.5.g1, alpha.5.g2)*1

Factor6 ~ c(NA, NA)*1 + c(alpha.6.g1, alpha.6.g2)*1")

syntax.time.bifactor[[4]] <- update(syntax.time.bifactor[[4]],

change.syntax = "g ~ c(NA, NA)*1 + c(alpha.1.g1, alpha.1.g2)*1

Factor1 ~ c(NA, NA)*1 + c(alpha.2.g1, alpha.2.g2)*1

Factor2 ~ c(NA, NA)*1 + c(alpha.3.g1, alpha.3.g2)*1")

#function for all MI tests – note that initially this was done step-by-step as more stringent comparisons cannot be made if less stringent comparisons are not reasonable

mi_test <- function(syntax, mod.cat, df.cat, group){

mod.config <- as.character(syntax[[1]])

fit.config <- cfa(mod.config, data = df.cat, group = group, parameterization = "theta")

mod.thresh <- as.character(syntax[[2]])

fit.thresh <- cfa(mod.thresh, data = df.cat, group = group, parameterization = "theta")

mod.metric <- as.character(syntax[[3]])

fit.metric <- cfa(mod.metric, data = df.cat, group = group, parameterization = "theta")

mod.scalar <- as.character(syntax[[4]])

fit.scalar <- cfa(mod.scalar, data = df.cat, group = group, parameterization = "theta")

out <- list(summary(compareFit(fit.config, fit.thresh)),

summary(compareFit(fit.thresh, fit.metric)),

summary(compareFit(fit.metric,fit.scalar)))

return(out)

}

out.age.cross <- mi_test(syntax.age.cross, nih6cross, recode_DSMage, "agegroup")

out.age.bi <- mi_test(syntax.age.bifactor, bifactor, recode_DSMage, "agegroup")

out.sex.cross <- mi_test(syntax.sex.cross, nih6cross, recode_DSMsex, "sex")

out.sex.bi <- mi_test(syntax.sex.bifactor, bifactor, recode_DSMsex, "sex")

out.time.cross <- mi_test(syntax.time.cross, nih6cross.sub, long_bl, "monthgrp") # omitting hearing vars zero variance

out.time.bi <- mi_test(syntax.time.bifactor, bifactor.sub, long_bl, "monthgrp") # omitting hearing vars zero variance.
